# COVID–19 Disease Dynamics in Germany: First Models and Parameter Identification

**DOI:** 10.1101/2020.04.23.20076992

**Authors:** Thomas Götz, Peter Heidrich

## Abstract

Since the end of 2019 an outbreak of a new strain of coronavirus, called SARS– CoV–2, is reported from China and later other parts of the world. Since January 21, WHO reports daily data on confirmed cases and deaths from both China and other countries [1]. The Johns Hopkins University [5] collects those data from various sources worldwide on a daily basis. For Germany, the Robert–Koch–Institute (RKI) also issues daily reports on the current number of infections and infection related fatal cases [2]. However, due to delays in the data collection, the data from RKI always lags behind those reported by Johns Hopkins. In this work we present an extended SEIR–model to describe the disease dynamics in Germany. The parameter values are identified by matching the model output to the officially reported cases. An additional parameter to capture the influence of unidentified cases is also included in the model.

There’s an evil virus that’s
threatening mankind […]
A menace to society
Iron Maiden, *Virus*, 1996.

## 1. Introduction

In December 2019, first cases of a novel *pneumonia of unknown cause* were reported from Wuhan, the seventh–largest city in China. In the meantime, these cases have been identified as infections with a novel strain of coronavirus, called SARS–CoV–2. Its genome sequence turned out to be 75– to 80–percent identical to the SARS–coronavirus, that caused a major outbreak in Asia in 2003. At the beginning of January 2020, the virus spread over mainland China and reached other provinces. Increased travel activities due to the Chinese new year festivities supported the expansion of the infection. From January 21 onwards, WHO’s daily situation reports contain the latest figures on confirmed cases and deaths, see [1].

The first COVID–19 case in Germany was reported in late January 2020 in a company close to Munich, Bavaria. Later cases were imported by travelers from China, Iran or Italy as well as tourists returning from ski holidays in the Austria and Italy. By 1 March 2020 more than 100 cases were reported in Germany and since than the number of cases began to rise exponentially. The first deaths were reported on 9 March 2020 [2, 6].

By 16 March 2020 the federal government introduced first measures to reduce the spread of the disease: Schools, kindergartens and universities were closed. On 22 March these measures were tightened by implementing a national curfew and contact ban. People are advised to stay at home, leaving only for work related activities, necessary shopping, medical treatment or sports. All this should not be done in groups of more than two persons if they do not belong to the same household [4].

Our work is based on the data reported by Johns Hopkins University [5]. We refrain from using the official data from the Robert–Koch–Institute [2], since they suffer from a delay by several days due to the more complicate way of aggregating those data.

## 2. Mathematical Model

To model the dynamics of the spread of COVID–19 incidences, we propose a hierarchy of SEIR–based models. For details regarding the original SIR– and SEIR–model we refer to classical works on mathematical epidemiology, e.g [7]. For our basic SEIRD–model, the total population of Germany with *N* ∼83.000.000 individuals is subdivided in to *susceptibles S, exposed E, infected I, recovered R* and *deaths D*. The susceptibles constitute the reservoir of persons that are not yet infected with SARS–CoV–2. After infection susceptible become exposed meaning that they already carry the virus but are not yet infectious. With a rate *θ* exposed individuals become infectious and transmit the virus with rate *β* to susceptibles. An infected individual loses infectivity with *γ* and has a probability *µ* of dying due to the disease [8]. Figure 2 shows the transmission structure. By *C* we denote all infected cases, independent of their current status. This artificial compartment is later on used to compare with the total number of registered cases reported by Johns Hopkins or RKI.

**Figure 1:**
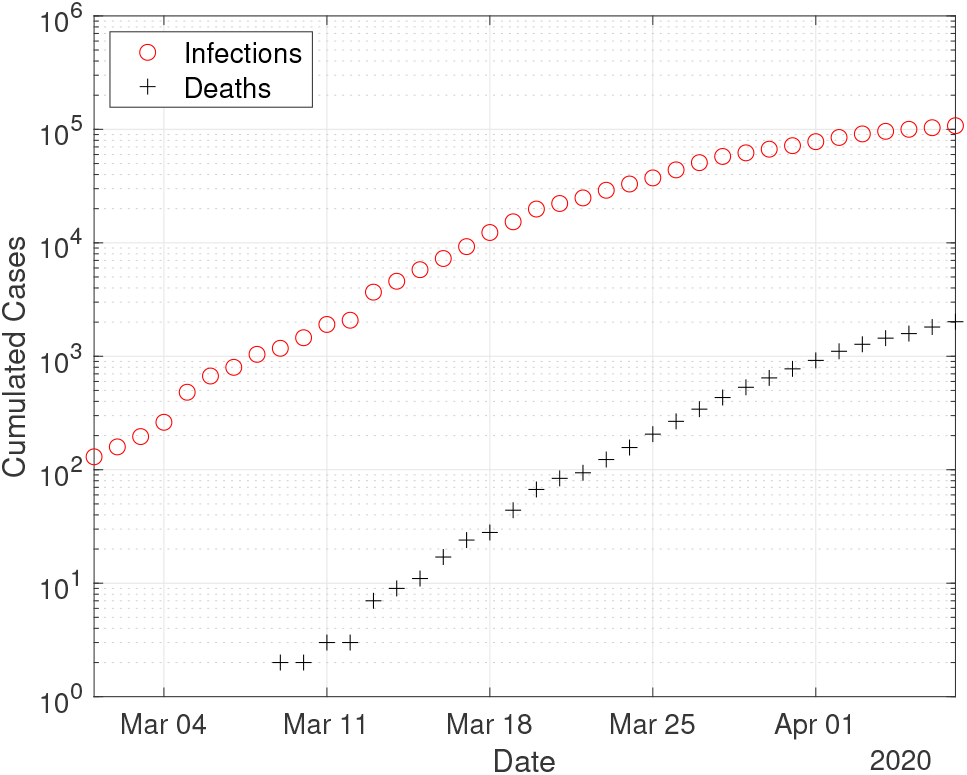
Case numbers in Germany from 1 March until 7 April 2020, as reported by Johns Hopkins University [5]. The initial time point is chosen as 1 March, since then the number of registered infections exceeds 100 cases.

**Figure 2:**
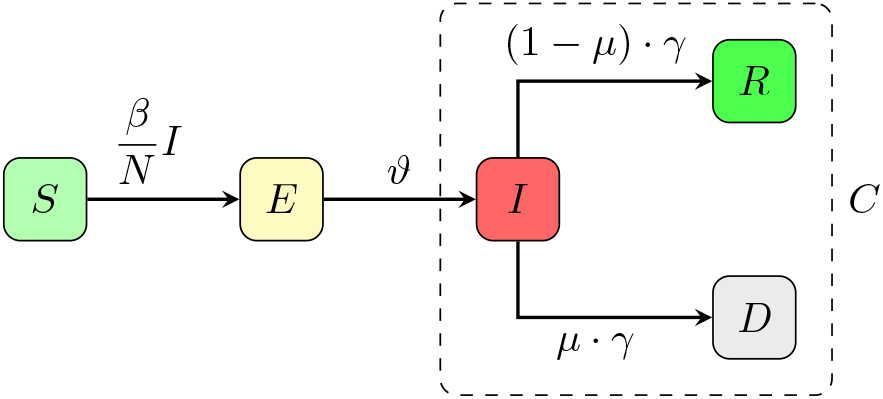
Transmission diagram for the basic SEIR–model (1). The artificial compartment C contains all infected cases, i.e. current active infections, recovered and deaths.

The resulting system of ordinary differential equations (ODE) for the above described SEIR–model reads as

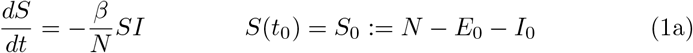

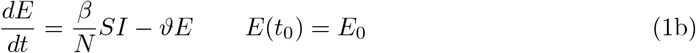

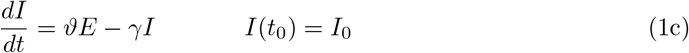

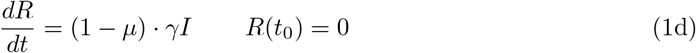

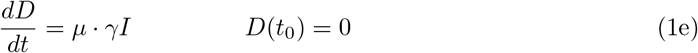

The starting time *t*_0_ is chosen as 1 March and the initial conditions for the recovered and dead compartment are assumed to be zero, since in Germany the first COVID–19 related death was recorded on 9 March. Also we may assume that the number of recovered individuals by 1 March is negligible.

In the sequel, we will also consider two refined versions of the above basic model.

At the onset of the disease, the numbers of exposed, infected, recovered and dead are still small and the number of susceptibles is approximately equal to the entire population *N*. In this setting, the *EI*–part of the model reduces to

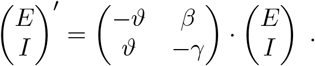

The maximal eigenvalue *λ* of this linear system determines the initial growth rate and is given by

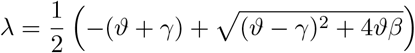

and the *doubling time T*_2_ equals

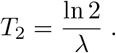

Figure 3 depicts the dependence of the doubling time on the transmission rate *β*. As of mid April, the doubling time in Germany is approximately 14 days compared to 2.5 days by mid March.

**Figure 3:**
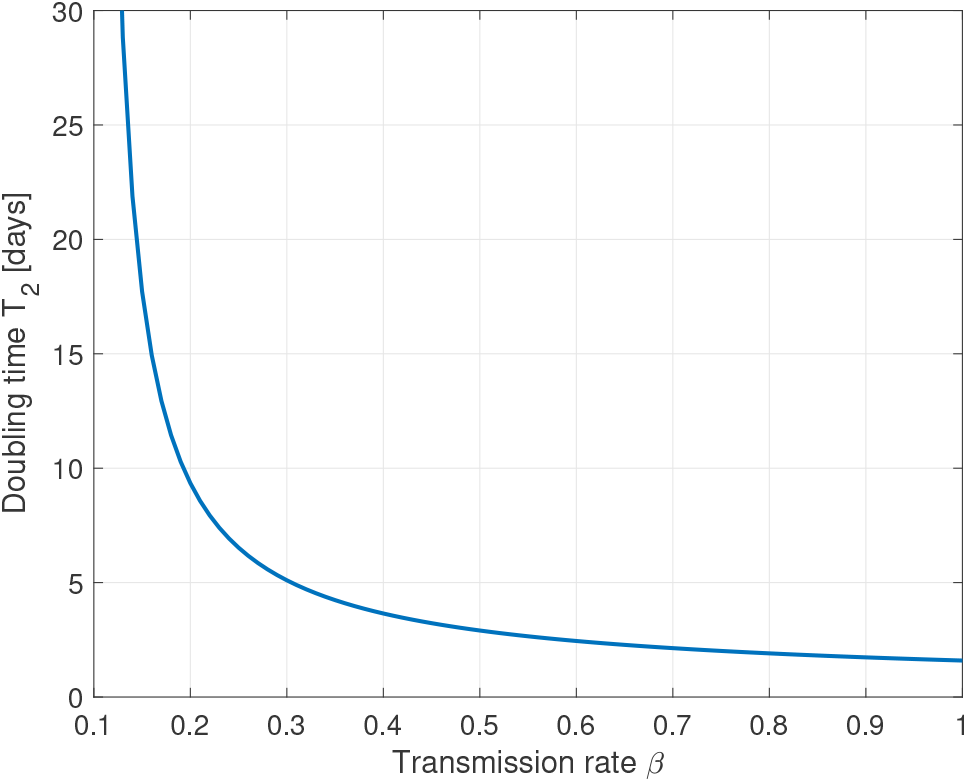
Plot of the doubling time *T*_2_ in days versus the transmission rate *β* for fixed values *θ* = 1*/*2 and *γ* = 1*/*10. A reduction of the transmission rate from *β* = 0.8 to *β* = 0.2 accounts for a slow down of the infection from doubling time 2 days to 10 days.

In the basic model (1), the transmission rate *β* is assumed to be fixed. The german state and federal governments introduced several measures to slow down the spread of the disease. Similar measures are nowadays taken in almost every country worldwide. From 16 March onwards schools, kindergartens and universities were closed and on 22 March a general contact ban was enforced in Germany. Both measures aim at reducing the transmission rate *β*.

To include this into the basic model (1), we also consider an alternative model for the transmission rate *β*: We assume *β* as a piecewise constant function on the time intervals prior to any measures, (until 15 March), after school closings (between 16 and 22 March) and after the contact ban (after March 22)

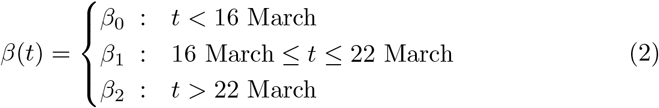

The resulting time–dependent SEIRD–model reads as

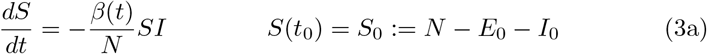

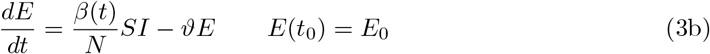

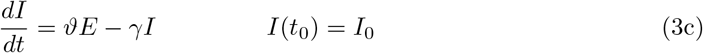

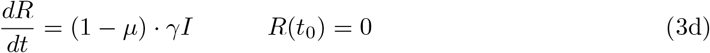

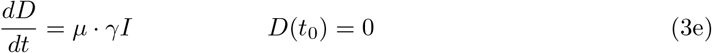

Setting *β* := *β*_0_ = *β*_1_ = *β*_2_, the time–dependent model reduces to the basic one. In order to validate our models and to identify the parameters involved therein, both the registered number of infections and the registered number of COVID–19 related deaths are important indications. The number of registered deaths is probably considerably more reliable, since the number of registered infections depends on the number of tests conducted and the dark figure of undetected, mostly asymptomatic cases, is assumed to be remarkably large [9]. We will discuss this point later in more detail. In the previous basic or time– dependent SEIRD–model, the actual increase of the disease related deaths 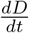 is assumed to be proportional to the current number of infected persons. The Robert–Koch–Institute specifies an average of 10 days between the onset of symptoms and admission to the intensive care unit [3]. Therefore, we assume *τ* = 14 for the time between the onset of infectiousness and death. In order to include this time lag into our model, we introduce a delay–term into the time–dependent model and obtain the final delayed time–dependent model:

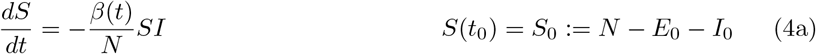

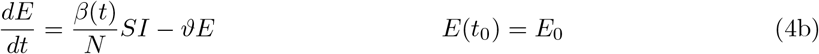

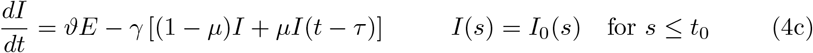

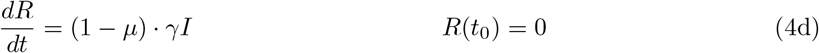

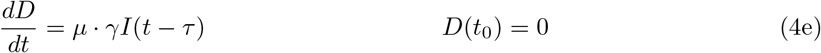

Note, that for solving this delay differential equation (DDE) we need an initial history of the infected compartment, i.e. values *I*_0_(*s*) for *t*_0_ *– τ ≤ s ≤ t*_0_.

In all the three models, the parameters *θ* = 1*/*2 [days^−1^], *γ* = 1*/*10 [days^−1^] are assume to be fixed and resemble a latency period of 2 days and a recovery period of 10 days, see [2, Situation report 31 March 2020].

The parameters in the transmission rate, i.e. *β*, or *β*_0_, *β*_1_, *β*_2_ the lethality *µ* and the initial values *E*_0_, *I*_0_ resp. the initial history *I*_0_(*s*) for the exposed and infected compartment are yet unknown to us. We will identify them together with the *detection rate δ* by matching the model output to the given data. The detection rate *δ* corresponds to the fraction of infected individuals which are positively tested for SARS–CoV–2 and hence appear in the official recordings. Various sources speculate that this detection rate is in the order of magnitude of 10–20% meaning that the true number of infected 5 – 10 times larger than the number published in the official statistics, see [9].

For the matching between the model output and the reported data we use a least–squares functional

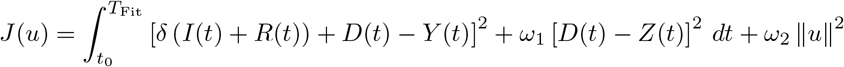

Here *u* = (*β, δ, µ, E*_0_, *I*_0_) resp. *u* = (*β*_0_, *β*_1_, *β*_2_, *δ, µ, E*_0_, *I*_0_) denote the unknown model parameters. Let *Y* (*t*) and *Z*(*t*) denote the data for the cumulated infected and dead cases at time *t* reported by Johns Hopkins University. The cumulated infected *Y*, i.e. total positive tests, are to be matched in the SEIRD–model to those individuals who had been infected until time *t*, i.e. the sum of the infected *I*, recovered *R* and deaths *D*. To account for the uncertainty in the *true* number of infected and recovered cases, we multiply both compartments by the detection rate *δ*, which is itself part of the parameters to be identified. For the deaths we assume no undetected cases. By *T*_Fit_ we denote the time horizon used for the comparison between the model and the data. The regularization term *ω*_2_ ‖ *u* ‖^2^ is included to ensure the convexity of the cost–functional. The weighting parameters *ω*_1_, *ω*_2_ *>* 0 allow to balance the contributions from the least squares error in the fatalities and from the size of the parameter values themselves to the least squares error in the infected cases. The relative weight for the infected cases is chosen to be equal to 1. The weight *ω*_1_ for the fatal cases allows to compensate the different order of magnitude between the infected cases and the fatal cases, typically *ω*_1_ ≃ 10–500. The weight *ω*_2_ is chosen small, such that the overall cost functional is still dominated by the least square fit between the model output and the given data.

The parameters *u*^*∗*^ themselves are obtained from minimization problem

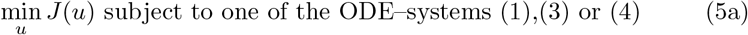

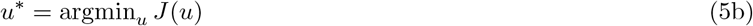

## 3. A few analytical considerations

Due to the absence of demographic terms, our basic model (1) does not allow other equilibria besides the trivial disease free equilibrium *X*^0^ = (*N*, 0, 0, 0, 0). Since we focus only on the short–time behavior of the epidemics, demographic terms are excluded and equilibria do not play any important role.

An important issue is the question of wether we can identify the detection rate and lethality during the take–off period of the epidemics? The only data available for parameter identification are the total number of registered cases *C* = *I* + *R* + *D* and the deaths *D*. The total registered cases heavily depend on the number of tests conducted. If a person is infected, but not tested, this person will not appear in the official statistics. Hence, there is a presumably large dark figure in the officially recorded data. Our model parameter *δ* takes this into account. The other, may be more reliable, available data are the recorded deaths. Here we may assume that *all* COVID–19 related deaths are diagnosed and hence there is no dark figure in the *D*–compartment. However, one scenario could be possible. A large dark figure in the entire cases, i.e. a small detection rate *δ* and a very small lethality could result in the same or at least similar observed data as a moderate or even small dark figure and hence large detection rate *δ* combined with a higher lethality rate. In that setting a simultaneous identification of both, the detection rate *δ* and the lethality *µ* could be difficult due to their counteracting effects.

In order to investigate this scenario, we consider the simultaneous effect of the detection rate *δ* scaling both the initial values of the *E* and *I* compartment to account for undetected cases together with a lethality *δµ*. Renoving the *S*– compartment by setting *S* = *N – E – I* − *R* − *D*, the basic SEIRD–system (1) reads as

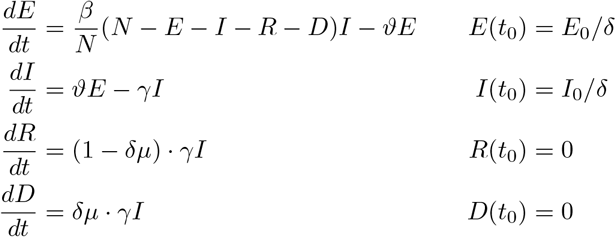

The sensitivities Σ_*E*_ := *∂*_*δ*_*E* and Σ_*I*_, Σ_*R*_, Σ_*D*_ of the solution with respect to the detection rate satisfy the system

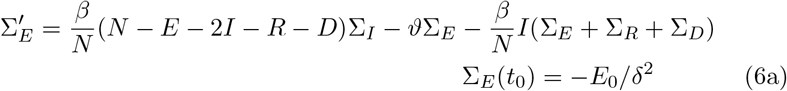

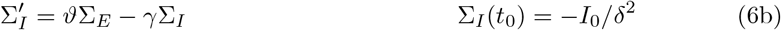

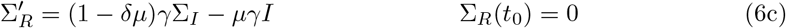

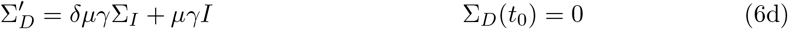

In Figure 4 we show the relative sensitivities Σ_*C*_*/C* and Σ_*D*_*/D* for detection rates *δ* = 0.1, 0.2 and 0.33. The chosen initial values are *E*_0_ = 150 and *I*_0_ = 100 (detected) cases at day 0. All other parameters resemble the assumed values for Germany. Note, that at the onset of the epidemics, i.e. in case of *δ* = 0.1 for *t* ≲ 30 and for *δ* = 0.2, 0.33 even for *t* ≲ 40, the sensitivities are very small and hence the solution of the SEIR–model is almost *independent* of the particular value of the detection rate *δ*. Hence *δ* cannot be identified from the observed data in a reliable manner.

**Figure 4:**
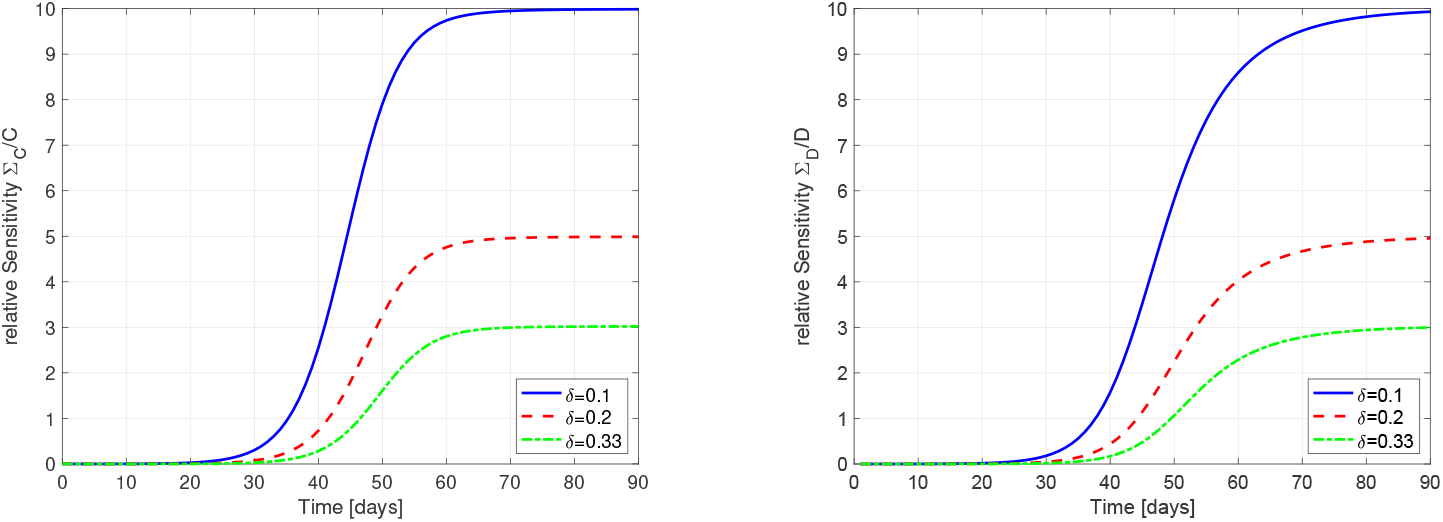
Relative Sensitivites of *C* (left) and *D* (right) with respect to the detection rate *δ* for *δ* = 0.1 (blue solid), *δ* = 0.2 (red dashed) and *δ* = 0.33 (green dash–dotted). At the onset of the epidemics, the sensitivities are extremely small, hence no reliable identification of *δ* is possible.

To illustrate these findings, we consider a linearization of a simplified SIR– model during the initial phase of the epidemics. We neglect the exposed compartment and assume that at the initial phase, the number of susceptibles is approximately equal to the entire population. Hence we get the linear system

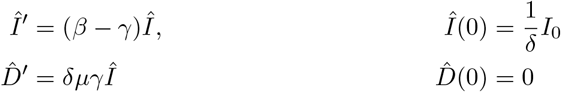

with the solution

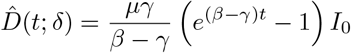

In this linearized setting, the approximation 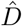 for the dead compartment is independent of the detection rate *δ*.

From the graphs in Figure 4 one can conclude, the a significant dependence of the detected or dead compartment *C* resp. *D* is given only after the initial take–off period of the epidemic. In the setting of Germany, this implies, that during the month of March a reliable identification to the detection rate might not be possible.

## 4. Adjoint Equations and Optimization

In order to solve the minimization problem (5), we use the adjoint equations, for details see [10, 12]. We introduce the Lagrangian

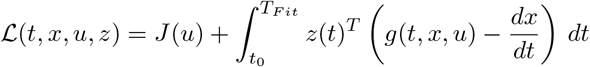

Here *z* = (*z*_*S*_, *z*_*E*_, *z*_*I*_, *z*_*R*_, *z*_*D*_) denotes the adjoint functions to the state variable *x* = (*S, E, I, R, D*) and *g*(*t, x, u*) denotes the right hand side of the ODE resp. DDE system. The gradient of ℒ with respect to the unknown parameters *u* is given by

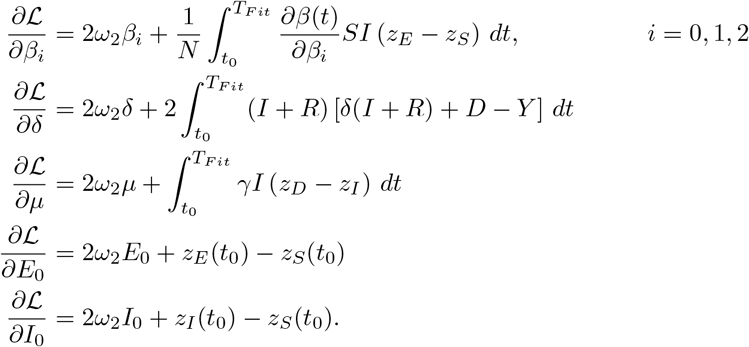

Note, that in the case *β* = *β*_0_ = *β*_1_ = *β*_2_ we have 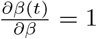 = 1. By adding the time delay, we obtain

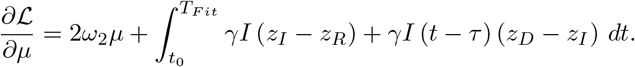

The adjoint system reads as

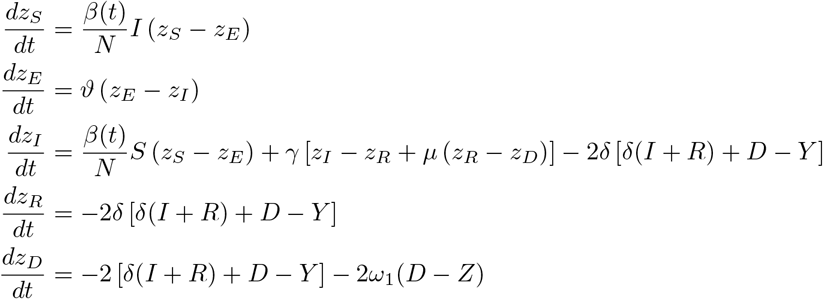

supplemented by the terminal condition (*z*_*S*_, *z*_*E*_, *z*_*I*_, *z*_*R*_, *z*_*D*_)(*T*_*F it*_) = 0. In the case of the time delay we receive

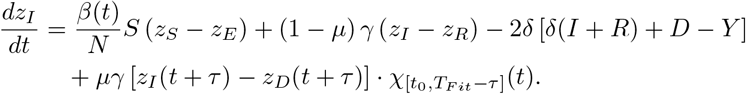

Here *χ*_[*a,b*]_(*t*) denotes the characteristic function of the interval [*a, b*], i.e. *χ*_[*a,b*]_(*t*) = 1 for *t ∈* [*a, b*] and = 0 otherwise.

To solve the optimization problem (5) numerically, we apply the Forward-Backward Sweep method [10] combined with a Quasi-Newton method *(BFGS)* [11].

In each iteration step the ODEs and DDEs of the state variables and adjoint equations are solved with Runge-Kutta methods before the corresponding gradient and direction of descent can be determined. The algorithm stops as soon as the termination condition ‖*J*(*u*_*k*+1_) − *J*(*u*_*k*_) ‖ *<* TOL is fulfilled.

As initial values we use *β* = *β*_0_ = *β*_1_ = *β*_2_ = 0.3 for the transmission rate.

This is justified by the fact that an average Basic Reproduction Number of about ℛ_0_ = 3 is assumed and in our basic model we have

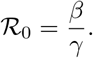

Epidemiologically, ℛ_0_ indicates the number of new infections an infected individual causes during the infectious period in an otherwise susceptible population. For the sake of simplicity, we assume the same starting value for *I*_0_ and *E*_0_. This corresponds to the value at the first data point of our measurement, i.e. 130 registered infected persons on 1st March. As already mentioned, we assume that for the recovered and deaths at this time *R*_0_ = *D*_0_ = 0 holds. The possible problems with the optimization of *δ* and *µ* were already mentioned in the previous section. To increase the probability of generating a global minimum, we use *n* = 1000 normally distributed start values for both parameters fulfilling *δ* ∼*𝒩* (0.25, 0.25^2^) and *µ* ∼*𝒩* (0.03, 0.03^2^) with *δ, µ >* 0. The algorithm selects the best result of these *n* data fits. The reason for this is the assumption that the proportion of detected cases is between 1 − 50% and the detected lethality between 1 − 6%. For the ratio of detected deaths 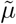 we have

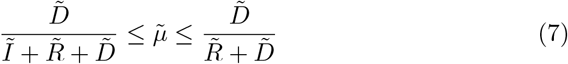

whereby, e.g. 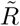 stands for the detected recovered at time *T*_*F it*_. The approach for this estimation can be found in [3]. It can be assumed that if the proportion of detected cases decreases, this also applies to lethality. The data records of Johns Hopkins University allow us to make this estimation in (7) [5].

In case of the time delay we choose as initial history for *s ∈* [*t*_0_ − *τ, t*_0_]

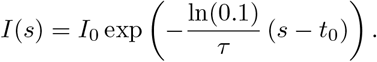

This is justified by the fact that the number of registered cases has increased tenfold during this period and we assume an exponential growth in this time span.

## 5. Simulation Results

To estimate the unknown parameters *u*, we match the data reported on a daily basis by Johns Hopkins [5] to our simulation results for a time period starting on 1 March.

The first results in Figure 5 show a parameter estimation using the basic model (1) and the time period before the onset of any containment measures, i.e. before the closing of schools on 16 March. We fitted the parameters *β, δ* and *µ* along with the initial values *E*_0_ and *I*_0_ over the time period 1 March to 16 March. The weight *ω*_2_ = 1 to keep the cost functional dominated by the two least square errors. The other weight is chosen as *ω*_1_ = 500 to compensate the significantly smaller value of the least square error in the fatal cases.

**Figure 5:**
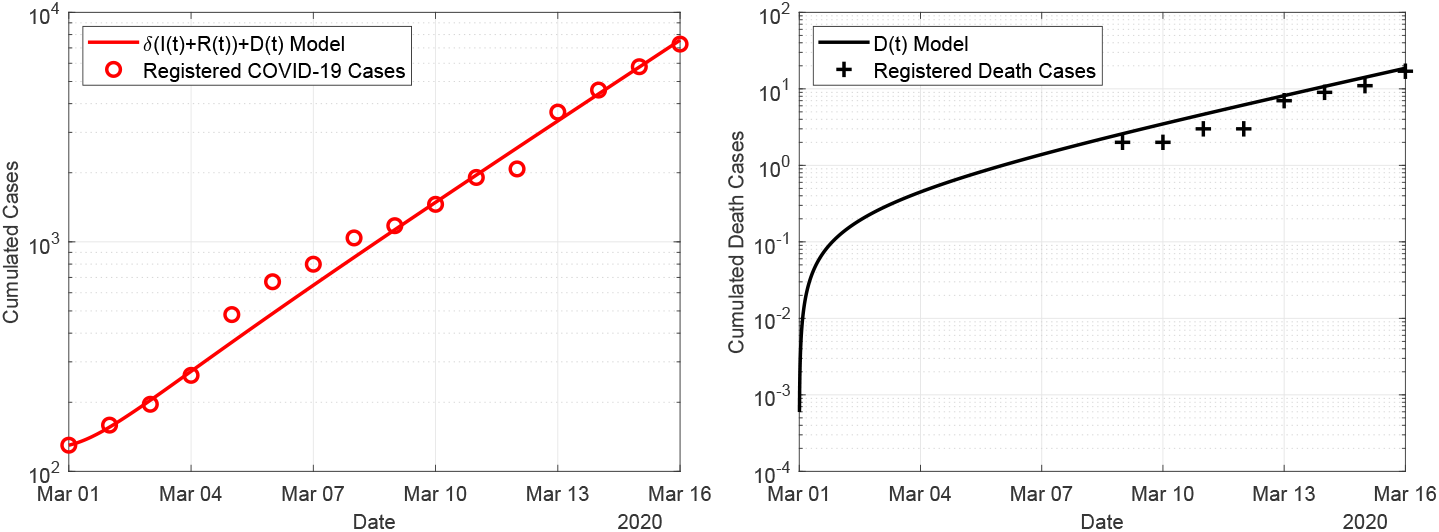
Fit of the basic model (1) to the data for the period 1 March to 16 March, i.e. before the onset of containment measures.

For the given time period of the fit, the model prediction and the observed data are in good accordance. The estimated parameter values are given in Table 1. The detection rate was estimated as *δ* = 0.37 implying that the *true* number of infections exceeds the registered cases by a factor 3. The transmission rate *β* = 0.57 accounts for a doubling time of 2.6 days at the initial, uncontrolled phase of the epidemic in Germany.

**Table 1:** Optimal parameter values for the three models (1), (3) and (4) obtained from the minimization problem (5).

| | Parameter | $\beta_0$ | $\delta$ | $\mu$ | $E_0 + I_0$ | $\beta_1$ | $\beta_2$ |
| --- | --- | --- | --- | --- | --- | --- | --- |
| Fit until | Model |  |  |  |  |  |  |
| 16.03.2020 | basic | 0.5656 | 0.372 | 0.0034 | 418 | — | — |
| 07.04.2020 | time-dep | 0.5232 | 0.308 | 0.0087 | 659 | 0.3561 | 0.1788 |
| 07.04.2020 | delay | 0.5532 | 0.202 | 0.0389 | 930 | 0.3578 | 0.1415 |

In Figure 6 we show the results obtained with the time–dependent model (3). In this case, the fitting period equals to the entire simulation period starting from 1 March to 7 April. The weights *ω*_1_, *ω*_2_ are identical to the previous simulation. The obtained transmission rate and according doubling times change from *β*_0_ = 0.5232 and *T*_2_(*β*_0_) = 2.8 days at the initial uncontrolled phase to *β*_2_ = 0.18 and *t*_2_(*β*_2_) = 11.4 days after the contact ban has been introduced. The effect of the contact ban effectively reduces the transmission rate by a factor of about 3 and significantly slows down the speed of the epidemics by increasing the doubling time by a factor 4.

**Figure 6:**
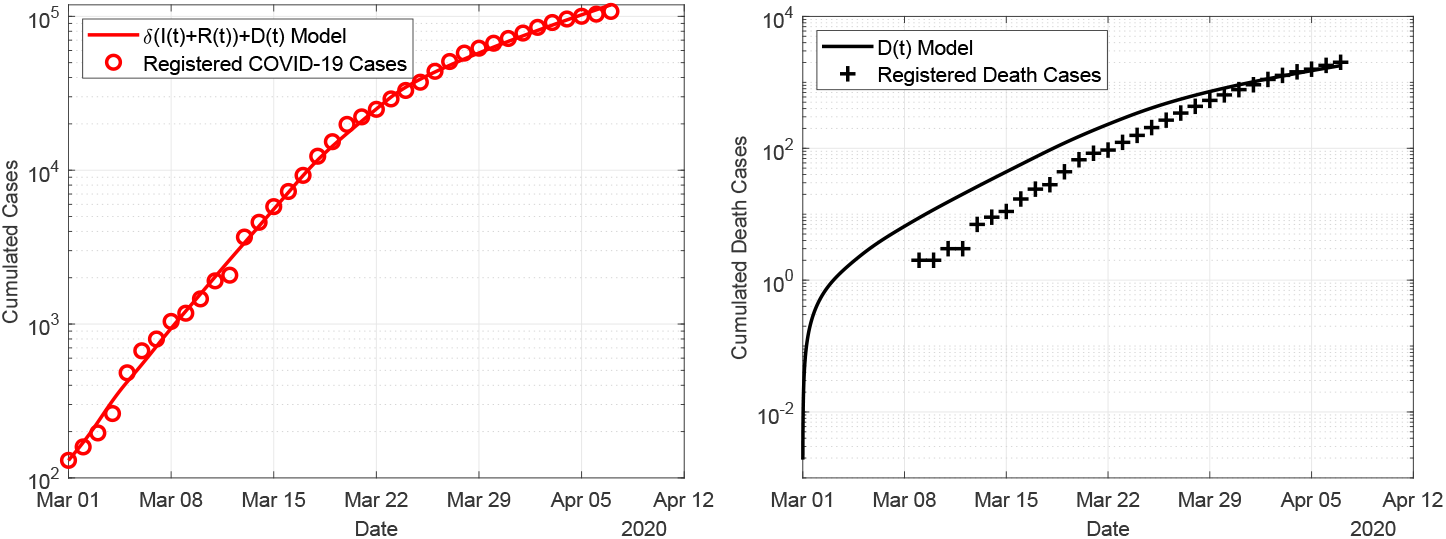
Fit of the time–dependent model (3) to the data for the period 1 March to 7 April.

In Figure 7 we show the result obtained with the delay model (4). For the delay model, we assume a delay of 14 days between entering the class of infected and death. Again, we show the simulation results compared to the reported cases for the infections and deaths. Excellent agreement is found between the model and the simulation for both, infections and deaths. Compared to the time–dependent model, shown in Figure 6, the delay model agrees better in particular for the fatal cases.

**Figure 7:**
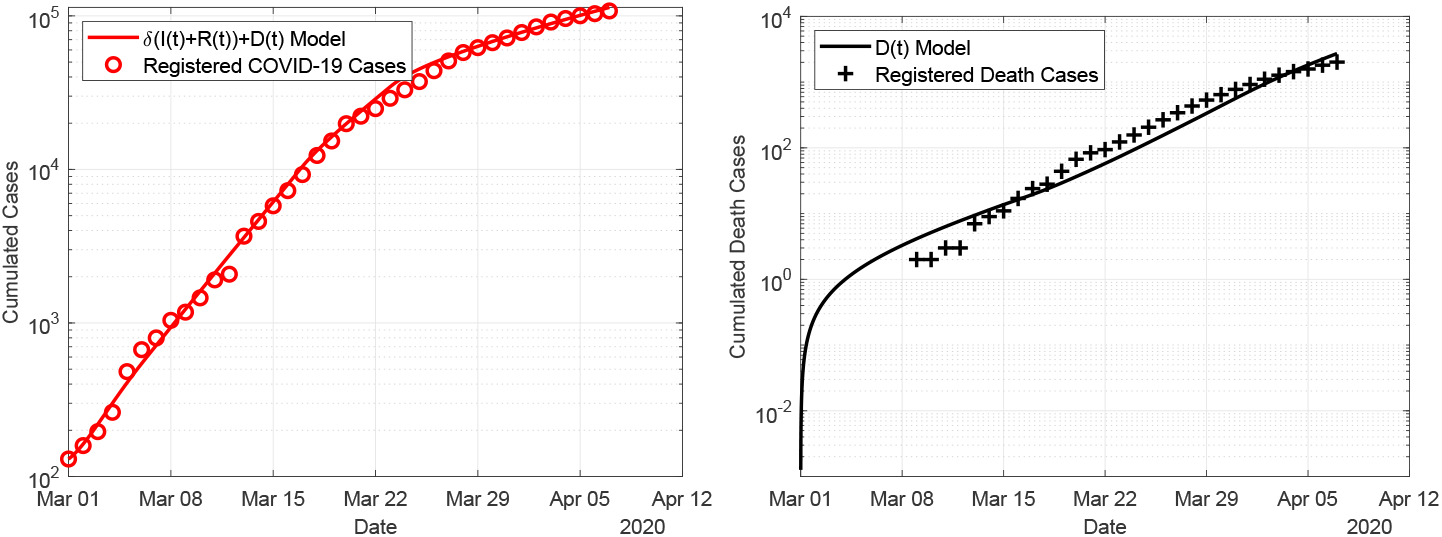
Fit of the delay model (4) to the data for the period 1 March to 7 April.

In the simulations the detection rate is found to be 20 − 40%, indicating that the *true* number of SARS–CoV–2 infections might be 3 − 5 times higher that the officially recorded data suggest. The lethality rate is found to be rather small, taking into account the large number of *true cases*.

Comparing the obtained values for the lethality, the value for the delay-model seems to be most realistic, since in this model we compare the fatal cases today to the infections that occurred two weeks ago. The two other models related the fatal cases of today to the infected cases *today*, hence to a significantly larger number. Therefore in these to models, the lethality rate seems to be smaller.

## 6. Conclusions and Outlook

We present three SIR–based models for describing the outbreak of the SARS–CoV–2 outbreak in Germany. Besides a standard SEIR–model, we consider an extension taking into account the effect of social distancing by a time–dependent reduction of the transmission rate. The third model introduces a delay–term to accurately describe the deaths depending on infected cases that occurred several days in the past. Comparing the simulation results to the data published by Johns Hopkins University allows an estimation of the unknown model parameters. Best results are obtained using the delay equation model. In this setting, we find a detection rate of about 20% and a lethality of about 4%. The social distancing measures were leading to an effective reduction of the transmission rate by a factor 4. That is, after the introduction of the measures roughly just 25% of the social contact compared to the initial period were leading to infections.

## Data Availability

All data used in the manuscript are open access.

